# Cost effectiveness of adopting a postoperative delirium risk prediction tool with non-pharmacological delirium prevention interventions for surgical patients

**DOI:** 10.1101/2024.08.07.24311487

**Authors:** Nicholas Graves, Soenke Boettger, Martin Zozmann, Maja Franziska, Reto Stocker

## Abstract

**Background:** Postoperative delirium arises among older surgical patients. Screening followed by prevention efforts are recommended. A risk prediction tool has been developed yet the performance and whether adoption is cost-effective are unknown.

**Objective:** To estimate the expected change to ‘total costs’ and ‘health benefits’ measured by quality adjusted life years from a decision to adopt PIPRA plus for screening purpose to find at-risk individuals who are then offered non-pharmacological interventions to reduce risks of postoperative delirium.

**Design:** Cost effectiveness modelling study that draws on a range of relevant data sources.

**Setting:** Swiss healthcare system.

**Subjects:** Surgical inpatients aged 60 or older, excluding cardiac and intracranial surgeries.

**Methods:** A decision tree model was used to capture the events likely to impact on cost and health outcomes. Information was harvested from a prospective before-after study and augmented with other relevant data sources. Probabilistic sensitivity analysis was undertaken to reveal the probability that adoption was cost effective against a stated maximum willingness to pay threshold for decision making in Switzerland.

**Results:** Patients in both phases of the study were similar. Costs were lower by 3075CHF (SD 997) per patient with the adoption of the risk screening tool and there was a modest gain to health benefits of 0.01 QALY (SD 0.026). There was a 100% probability that adoption would be cost saving and a 91% probability that adoption would be cost-effective.

**Conclusions:** We provide early-stage evidence that a decision to adopt the risk screening tool and offer risk reducing interventions will be cost-effective.

**Key points:** Many surgical patients suffer from post operative delirium.

Screening and early intervention can reduce risks and improve outcomes.

It is important to establish whether screening and early intervention is cost effective.

## Introduction

Postoperative delirium (POD) is a frequent and potentially serious adverse event that typically affects patients over 65 years [1]. Characterised by a sudden change to mental function it causes individuals to be disoriented, confused, and agitated. Hospital stay is often extended [2] and there is increased morbidity and mortality [3]. While some patients recover, the risks of dementia are increased [4] with dementia patients showing faster than normal functional decline and increased chances of being admitted to a long term care home [5].

Professional groups recommend patients at risk of POD be identified and offered prevention strategies [6, 7]. Multicomponent prevention interventions [8, 9] could prevent up to 40% of identified POD. Recognised non-pharmacological interventions include mobilising the patient, ensuring uninterrupted sleep, good hydration, prevention of healthcare associated infection, pain control and avoiding certain drugs that might cause delirium.

A risk prediction tool called ‘Pre-Interventional Preventive Risk Assessment’ or PIPRA [10, 11] has been developed for use by health care professionals, to identify patients over 60 years of age at risk of developing POD. Patients then receive prevention focused nursing-led interventions to reduce risks of POD. Implementing PIPRA into hospitals will increase costs through additional perioperative screening, data processing and provision of interventions. The potential exists for healthcare organisations to save resources by reducing the incidence of POD, shortening lengths of stay in hospital, increasing discharges home rather than to a facility, reducing risks of death and achieving better health related quality of life.

The aim of this cost-effectiveness modelling study is to estimate the expected change to ‘total costs’ and ‘health benefits’ measured by quality adjusted life years (QALYs), from a decision to adopt PIPRA into a hospital workflow. Specifically, there is screening of surgical inpatients aged 60 or older with PIPRA, excluding cardiac and intracranial surgeries, and the provision of targeted non-pharmacological prevention interventions to an identified at-risk group. The main outcomes are changes to total costs and QALYs. This cost-effectiveness modelling study adheres to the CHEERS reporting guideline [12].

The relevance for decision-making in practice is that preventive interventions and routine delirium screening are not routinely implemented in hospitals, and approximately 70% of POD cases remain undiagnosed [13]. Substantial economic benefits could arise from the use of an effective screening tool that leads to effective interventions.

## Methods

### Study Design

We use data from a quality improvement project evaluated using a before-after design [14] and completed in a 335-bed Swiss private hospital. The methods and detailed findings from this study have been reported [28]. The before-after study enrolled eligible surgical admissions from May 1^st^ to June 30^th^ 2023, aged 60 or over, and not admitted for cardiac or intra-cranial surgery.

The first phase of the study was to establish a “control” group from May 1^st^ to May 22^nd^. During this time, staff were educated on POD and the importance of regular delirium assessment and delirium treatment. Nurses were required to perform delirium screening three times a day, using the delirium observation screening scale (DOSS) [15].

The second phase, or “intervention”, ran from May 23^rd^ to June 30^th^ and included implementation of the PIPRA POD risk prediction tool to identify the at-risk group, and the subsequent application of targeted non-pharmacological preventive measures including history-taking upon admission, encouragement of family involvement, empathetic communication, daily assessment of whether a urinary and intravenous catheter is needed, early and regular mobilisation, multimodal pain management, and enhancement of sleep-wake patterns, such as exposure to daylight and avoidance of naps during daytime, and minimising light, noise and patient care during night-time. The nurses were also asked to orient the patients regularly, through communication and provision of pictures of relatives, personal items, hearing aids, glasses, and to communicate a structured daily schedule, supported by the provision of large clocks, calendars, and whiteboards. For both phases of the study information on the amount of nursing time and lengths of stay in a ward bed and an ICU bed were collected.

The perspective for this analysis is healthcare sector and we include cost outcomes for acute care, nursing homes and rehabilitation facilities. We exclude private costs incurred by patients and families due to a lack of reliable information. The time horizon for costs was <12 months. Health outcomes accounted for a normal life expectancy, and future health benefits were discounted at 3%.

### Cost effectiveness model

We developed a decision tree model to capture the events likely to impact on ‘cost’ and ‘health outcomes’ related to a decision to adopt the PIPRA POD screening tool into a hospital workflow, see Figure 1.

**Figure 1.**
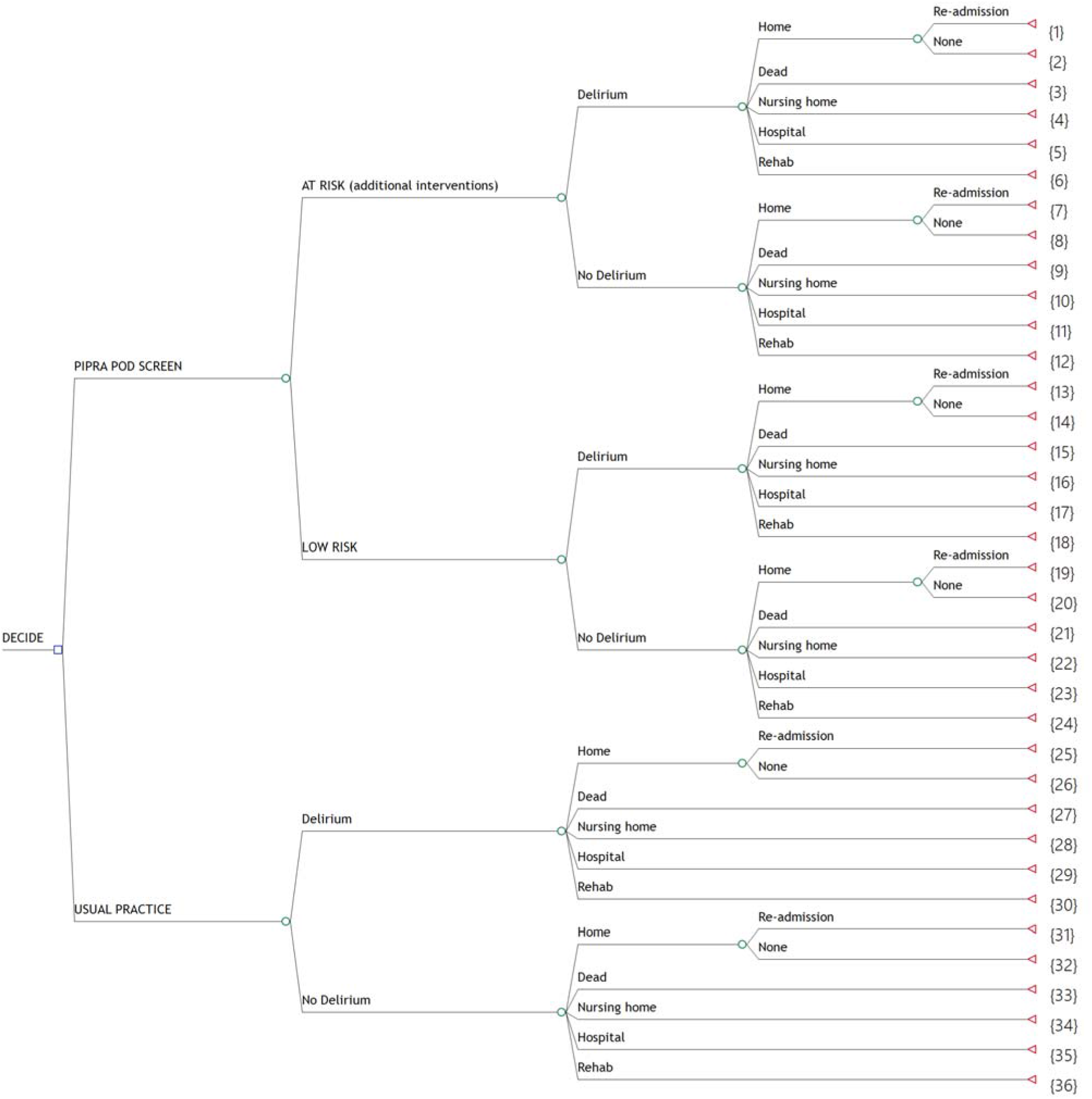
Decision Tree used to evaluate the cost-effectiveness of a decision to adopt PIPRA.

Under the ‘PIPRA POD SCREEN’ option patients were either classified ‘AT RISK,’ and received the relevant preventive measures, or, were classified as ‘LOW RISK’ and no extra measures were taken beyond standard of care. All patients were screened for POD and a clinical diagnosis of ‘POD’ was made if patients had at least one DOSS value of 3 or above during their hospital stay or had a formal clinical diagnosis of delirium. All patients have some probability of being discharged to home, nursing home, hospital, or a rehabilitation facility, or dying and this varied by their delirium status. Those discharged home face a further probability of being re-admitted to an acute hospital within 30 days. Under the ‘USUAL PRACTICE’ option, some patients were diagnosed with POD and all patients faced probabilities of the competing discharge options listed above. The total costs arising from all the events on the 36 unique branches were estimated at the terminal nodes marked with triangles. The health outcomes, measured by quality adjusted life years (QALYs) were also summarised at the terminal nodes. The probabilities of events were included at the circles and were used to calculate the expected costs and QALYs, which are finally summed at the ‘DECIDE’ node at the far left of the tree for the ‘PIPRA POD SCREEN’ and the ‘USUAL PRACTICE’ options. This enabled evaluation of the expected incremental change to ‘total costs’ and ‘health benefits’ from a decision to adopt PIPRA into the hospital workflow.

### Evidence synthesis for costs, QALYs and probabilities

The duration of a re-admission within 30 days of discharge from hospital was taken from a US nationwide in-hospital and readmission database for years 2010–2015 [16]. The costs per day for a nursing home were reported by the authors of a Swiss-based open two-phase randomized controlled trial at three nursing homes [17]. We assumed that only 6 months of the stay in a nursing home was related to the presence of POD, and beyond that time, the patient recovers and can return home. The amount of nursing time and lengths of stay in a ward and ICU bed(s) were taken from the before-after study completed in the Swiss private hospital [28]. The cost for a ward bed day was taken from a study of methicillin-resistant *Staphylococcus aureus* at a Swiss University hospital [18]; as this was published in 2013, the costs were adjusted to 2024 prices using 3% inflation. The cost for an ICU day was taken from a study of severe sepsis in Swiss hospitals [19]; as this was published in 2004, the costs were adjusted to 2024 prices using 3% inflation. The cost of nursing time was taken from a recruitment website that reported salary costs for nursing positions in Switzerland [20]. A charge of 25CHF was made for each patient being screened. See Appendix 1 for all cost parameters used.

For the estimation of QALYs, the years of life lost from a death was estimated as the difference between the life expectancy of Swiss males (85.57 years) and Swiss females (88.1 years) [21], and the mean age of patients enrolled in the interrupted time series study completed in the Swiss private hospital; this was 72.08 years for males and 72.29 for females. The proportion of males in the study was 54%. Future years of life were discounted by 3% per year to account for time preferences. Preference based health utility weights for relevant health states were estimated by Young *et al*. [22]. They mapped SF36 data using an algorithm [23] to a preference based measure using information collected from a study of 115 Swedish patients for which the aim was to compare change in cognitive function and health-related quality of life 6 months after hip surgery. In that study, 32 patients became delirious during their hospital stay [24]. See Appendix 2 for the health utility parameters used.

The probability of a ‘AT RISK’ designation under ‘PIPRA POD SCREEN’ and probability of ‘Delirium’ outcomes were estimated from the before-after study [28]. The probabilities of being discharged to the competing locations shown in the decision tree model were estimated from a pragmatic perspective cohort study of consecutive admissions to a large healthcare system in Switzerland [25]. This study reported relevant discharge destinations for 27,026 consecutive adults with length of stay of at least 24 hours in an acute hospital for those with and without delirium. The probability of being re-admitted within 30 days was taken from a study of 453 consecutive (≥65years old) patients undergoing spine surgery for whom a proportion had delirium [26]. See Appendix 3 for all parameters that update the probabilities used in the model.

### Model Evaluation and Assessment of Uncertainty

Clinical experts validated the structure of the decision tree familiar with the Swiss health care system. Model parameters were fitted to Beta, Dirichlet, Gamma, Uniform, and Normal distributions. Three thousand Monte Carlo samples were taken from the uncertain parameters to estimate a joint distribution of the incremental change to costs and QALY outcomes from a decision to adopt PIPRA into a hospital workflow. To interpret the results for decision-making, we assumed a decision-maker is willing to pay 100,000 CHF for a marginal QALY [27].

### Scenario Analysis

Three additional scenarios were modelled. The first assumed that the cost of a ward day is less than 3,715 CHF. The original estimate came from a university teaching hospital and might reflect higher overheads and operating expenses than is typical in smaller Swiss hospitals. Hence a lower estimate of 1,400 CHF was used in this scenario. The second additional scenario was that we assumed that only 6 months or 182.5 days of the stay in a nursing home was related to the presence of POD. This implies the patient does not recover, becomes demented and spends the rest of their life in a nursing home, incurring worse health outcomes and additional costs. For the final additional scenario, we excluded non-hospital costs and estimated the outcomes from the perspective of the hospital-level decision-maker only.

## Results

The characteristics of the patients in both phases are shown in Table 1, which is reproduced from the primary paper reporting clinical outcomes [28].

**Table 1.** Description of included subjects and outcomes.

|  | Overall | Phase 1<br>(control) | Phase 2<br>(intervention) | p-<br>value | Missing |
| --- | --- | --- | --- | --- | --- |
| Number of patients | 866 | 299 | 567 |  |  |
| Age in years [range] | 72 [66, 78] | 72 [66, 77] | 72 [66, 78] | 0.86 | 0.0 |
| Female (%) | 399 (46.1) | 129 (43.1) | 270 (47.6) | 0.24 | 0.0 |
| BMI [range] | 25.14 [22.73, 28.01] | 25.04 [22.95, 28.03] | 25.17 [22.52, 28.00] | 0.62 | 5.0 |
| Cognitive impairment (%) | 24 (3.7) | 9 (4.2) | 15 (3.5) | 0.81 | 25.2 |
| History of delirium (%) | 24 (3.7) | 5 (2.3) | 19 (4.4) | 0.27 | 24.5 |
| SPI [range] | 38 [33, 40] | 38 [33, 40] | 39 [34, 40] | 0.77 | 5.0 |
| No. Medications<sup>a</sup> [range] | 4 [2, 7] | 4 [2, 6] | 4 [3, 7] | 0.14 | 21.2 |
| log(CRP) | 0.97 [-0.69, 2.81] | 0.81 [-0.36, 2.47] | 0.99 [-0.69, 3.05] | 0.98 | 72.5 |
| Surgical risk<sup>b</sup> (%) |  |  |  | 0.12 | 0.1 |
| 1 | 253 (29.2) | 75 (25.1) | 178 (31.4) |  |  |
| 2 | 565 (65.3) | 205 (68.6) | 360 (63.6) |  |  |
| 3 | 47 (5.4) | 19 (6.4) | 28 (4.9) |  |  |
| Delirium risk<sup>c</sup> [range] | 0.07 [0.04, 0.16] | 0.07 [0.04, 0.16] | 0.08 [0.03, 0.16] | 0.91 | 0.0 |

Unless otherwise indicated, data shown are n (%-percentage of total) or median [IQR-interquartile range]. BMI: body mass index, SPI: "Selbstpflegeindex" self-care index, CRP: C-reactive protein, SD: standard deviation. <sup>a</sup>Preoperative, <sup>b</sup>cardiac risk for non-cardiac surgery, <sup>c</sup>PIPRA risk score

There are few differences between the patients in each phase, and none of the differences are statistically significant. We note that cognitive impairment is slightly more common in the Phase 1 control patients, history of delirium is more frequent in the Phase 2 intervention patients, C-reactive protein scores are greater in Phase 2 and more patients in Phase 2 have a Surgical risk of 1.

From those screened during the Phase 2 intervention period, 40.91% were found to be at-risk of developing POD and they received additional non-pharmacological interventions to address those risks. The observed effect of this was positive but modest, with the probability of delirium for the controls found to be 12.38% and 11.11% for the intervention patients. For those who had delirium, the probability of being discharged to their own home was 49.74% as compared to 91.66% for those without delirium. The probabilities of dying were 11.16% for delirium and 0.29% for non-delirium. The probability of being discharged to a nursing home, another hospital or a rehabilitation centre was lower for the non-delirium patients, see Table 2 for all results.

**Table 2.**
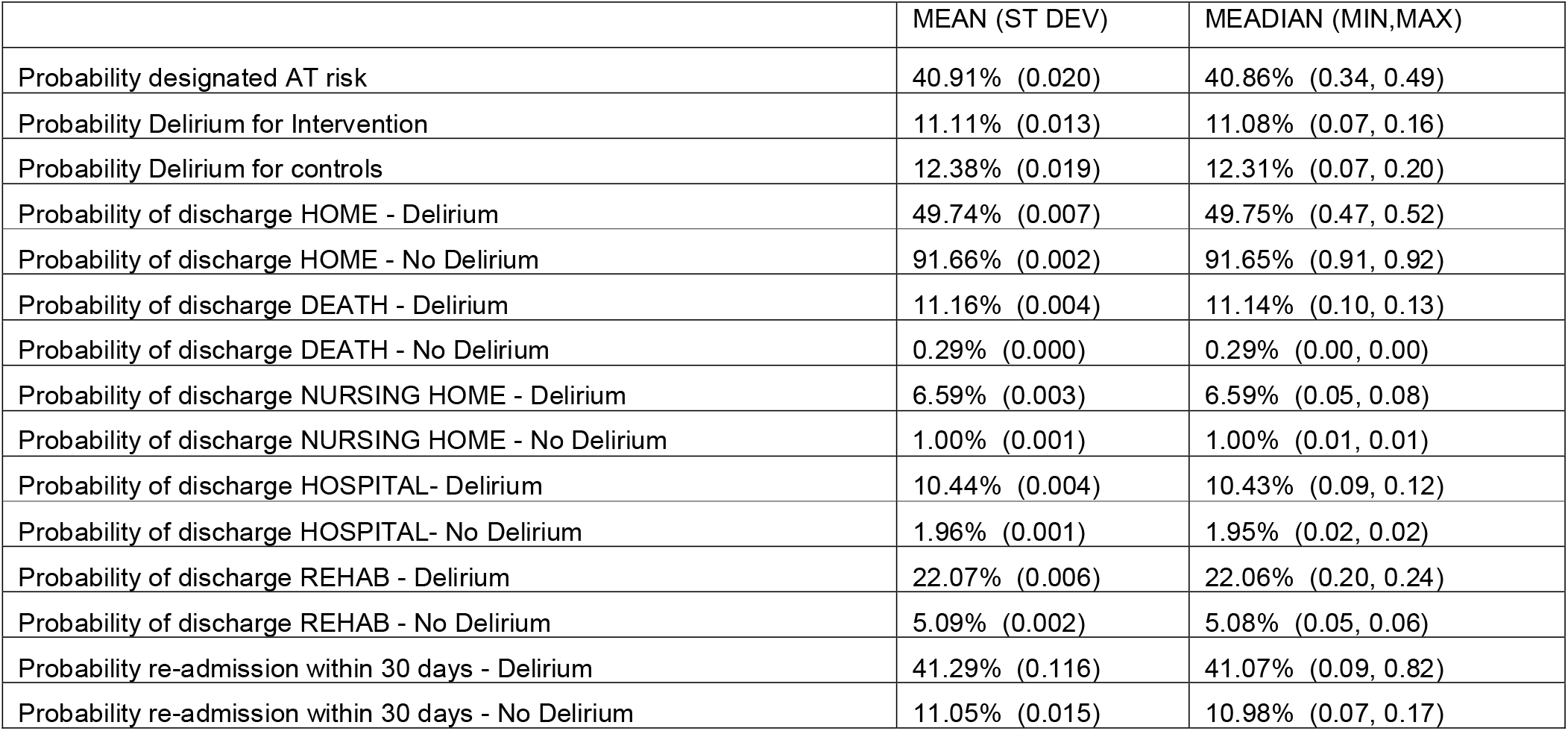
Modelled outcomes from the analysis of the decision tree.

The impact on costs for the intervention group was meaningful, while the impact on QALYS, although positive, was modest. Total costs were reduced by 3,075CHF per patient for the intervention group and QALYS were increased by 0.01, see Table 3.

**Table 3.** Modelled outcomes from Analysis of Decision Tree.

| *PRIMARY RESULTS* | *MEAN (ST DEV)* | *MEDIAN (MIN,MAX)* |
| --- | --- | --- |
| Total costs - CONTROL (CHF) | 24,622 (890) | 24,586 (21,993, 27,504) |
| Total costs - INTERVENTION (CHF) | 21,547 (753) | 21,550 (18,974, 24,476) |
| INCREMENTAL COST (CHF) | -3,075 (997) | -3,090 (-6,975, 0,022) |
| QALYS – CONTROL | 6.62 (0.022) | 6.63 (6.54, 6.69) |
| QALYS - INTERVENTION | 6.64 (0.016) | 6.64 (6.57, 6.68) |
| INCREMENTAL QALY (CHF) | 0.01 (0.026) | 0.01 (0, 0.11) |
| *HOSPITAL COSTS ONLY* | *MEAN (ST DEV)* | *MEDIAN (MIN,MAX)* |
| Total costs - CONTROL (CHF) | 20,437 (686) | 20,427 (18,378, 23,073) |
| Total costs - INTERVENTION (CHF) | 17,952 (596) | 17,944 (15,764, 20,385) |
| INCREMENTAL COST (CHF) | -2,485 (890) | -2,487 (-5,944, 0,928) |
| QALYS - CONTROL | 6.62 (0.022) | 6.62 (6.53, 6.68) |
| QALYS - INTERVENTION | 6.64 (0.015) | 6.64 (6.58, 6.69) |
| INCREMENTAL QALY (CHF) | 0.01 (0.026) | 0.01 (0, 0.13) |

Decision uncertainty shown in Figure 2 was negligible, with a 100% probability that adoption would be cost saving, and an 91% probability that adoption would be cost-effective at a threshold value of 100,000 CHF per QALY gained, shown by the red line.

**Figure 2.**
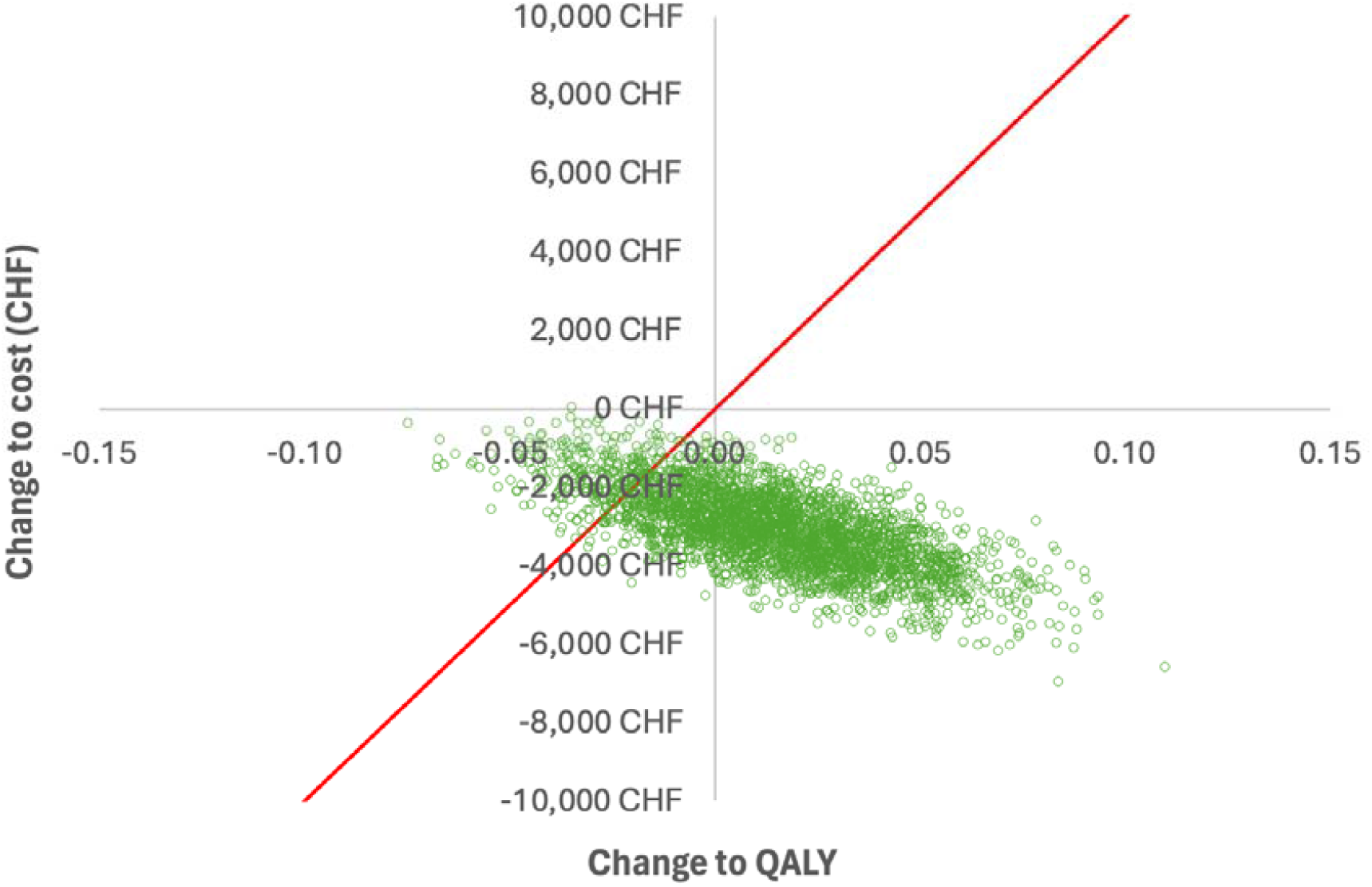
Finding from cost-effectiveness analysis showing decision uncertainty

The first additional scenario, where costs per acute bed day were reduced to 1,400 CHF, had almost no impact on the results. The second scenario, where we assumed the patient does not recover and spends the rest of their life in a nursing home, incurring lesser health outcomes and extra costs, would clearly strengthen the finding in support of cost-effectiveness. The final scenario of ‘hospital costs’ only, revealed the mean savings were increased to 2,485CHF per patient, see Table 3.

## Discussion

The findings of this cost-effectiveness study conducted in a Swiss setting suggest reasonable cost savings to health services and small gains to health outcomes. For the next 10,000 eligible patients, the adoption of PIPRA POD risk prediction and targeted non-pharmacological interventions, would result in resource saving to the healthcare system valued at 30M CHF, and for the hospital sector only, these savings are expected to be 25M CHF. The effect of implementing PIPRA on incidence rates of POD was modest, with only a 1.3% reduction. However, there had been large efforts made in the Phase 1 control period to educate healthcare workers on POD recognition and the importance of regular assessment, which could have reduced the risks of POD prior to PIPRA being implemented.

There are limitations to this analysis and these initial findings should be interpreted with caution. The probabilities of having POD arose from a small observational study conducted in a single private hospital, and not a randomised design, hence we cannot be certain that the reduction in POD was a causal effect. The observed cost savings cannot be thought of as cash savings, as most of the costs of healthcare services are fixed and cannot be escaped in the short term [29]. Instead, resources such as bed days, nursing time and capacity in nursing homes are released, and these have positive economic value. The parameters used to describe the health utilities were assumed to be fixed for the model, as the information required to make them probabilistic was not available. It is however unlikely that this omission will change the decision-making conclusions. We assumed a marginal QALY was worth 100,000 CHF to decision makers, but this assumption was not based upon empirical data. Ideally, the marginal productivity of health spending is used to set a threshold [30], but this was not available for the Swiss setting. Again though, due to the large probability that adoption is cost-saving, this assumption is unlikely to change the conclusions for organisational decision-making.

## Conclusions

We provide preliminary evidence that a decision to adopt PIPRA POD risk prediction, together with targeted non-pharmacological preventive strategies, will save healthcare and hospital costs, and will be cost-effective. Ideally, this intervention will be evaluated further in a randomised trial and in multiple settings.

## Data Availability

All data produced in the present work are contained in the manuscript

## Appendices

**Appendix 1.** Cost Parameters for Decision Tree Model

|  | *Estimate* | *Distribution* |
| --- | --- | --- |
| Duration of re-admission for delirium in days | 7.7 | Uniform (6, 9) |
| Duration of re-admission for non-delirium in days | 6.6 | Uniform (5, 8) |
| Cost per day for Nursing Home stay (CHF) | 80 | Uniform (80, 175) |
| Cost per ward Day (CHF) | 3715 | Fixed |
| Cost per ICU Day (CHF) | 5859 | Fixed |
| Cost per hour of nursing time (CHF) | 42 | Fixed |
| Cost per patient for PIPRA screen (CHF) | 25 | Fixed |
| *Nursing Time in minutes for study patients* | *Mean (s.e.)* | *Distribution* |
| usual practice 'delirium' dead | 1554 (8.9) | Gamma (30517, 0.05) |
| usual practice 'delirium' alive | 3154 (98.6) | Gamma (1023, 3.08) |
| usual practice 'no delirium' dead | 420 | Fixed |
| usual practice 'no delirium' alive | 1240 (49.0) | Gamma (640, 1.94) |
| at risk 'delirium' dead | 4649 (40.0) | Gamma (13536, 0.34) |
| at risk 'delirium' alive | 4105 (161.7) | Gamma (644, 6.37) |
| at risk 'no delirium' dead | 1357 (51.5) | Gamma (693, 1.96) |
| at risk 'no delirium' alive | 1438 (49.8) | Gamma (834, 1.72) |
| low risk 'delirium' alive | 1488 (27.8) | Gamma (2865, 0.52) |
| low risk 'no delirium' alive | 914 (23.1) | Gamma (1564, 0.58) |
| *ICU Hours used for study patients* | *Mean (s.e.)* | *Distribution* |
| usual practice 'delirium' dead | 24.8 (1.2) | Normal |
| usual practice 'delirium' alive | 11.8 (1.2) | Normal |
| usual practice 'no delirium' alive | 1.4 (0.3) | Normal |
| at risk 'delirium' dead | 96.5 (2.0) | Normal |
| at risk 'delirium' alive | 14.1 (1.2) | Normal |
| at risk 'no delirium' dead | 2.5 (0.5) | Normal |
| at risk 'no delirium' alive | 3.7 (13.5) | Normal |
| low risk 'no delirium' alive | 2.2 (0.7) | Normal |
| *Length of stay in days for study patients* | *Mean (s.e.)* | *Distribution* |
| usual practice 'delirium' dead | 4.2 (0.06) | Gamma (5584, 0.001) |
| usual practice 'delirium' alive | 10.6 (0.26) | Gamma (1679, 0.006) |
| usual practice 'no delirium' dead | 0.9 | Fixed |
| usual practice 'no delirium' alive | 4.5 (0.15) | Gamma (873, 0.005) |
| at risk 'delirium' dead | 8.4 (0.05) | Gamma (30167, 0.0003) |
| at risk 'delirium' alive | 10.1 (0.27) | Gamma (1373, 0.007) |
| at risk 'no delirium' dead | 2.5 (0.07) | Gamma (1149, 0.002) |
| at risk 'no delirium' alive | 5.0 (0.15) | Gamma (1106, 0.004) |
| low risk 'delirium' alive | 6.0 (0.12) | Gamma (2378, 0.003) |
| low risk 'no delirium' alive | 3.6 (0.09) | Gamma (1748, 0.002) |

**Appendix 2.**
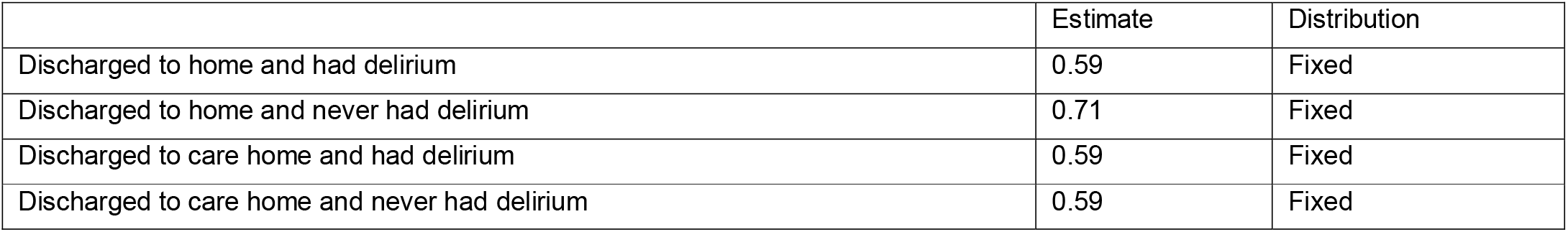
Health Utility Parameters for Decision Tree Model

|  | Estimate | Distribution |
| --- | --- | --- |
| Discharged to home and had delirium | 0.59 | Fixed |
| Discharged to home and never had delirium | 0.71 | Fixed |
| Discharged to care home and had delirium | 0.59 | Fixed |
| Discharged to care home and never had delirium | 0.59 | Fixed |

**Appendix 3.** Probabilities used for Decision Tree Model

|  | Estimate | Distribution |
| --- | --- | --- |
| Probability of AT risk designation | 40.92% | Beta (232, 335) |
| Probability of Delirium AT Risk | 22.84% | Beta (53, 179) |
| Probability of Delirium LOW Risk | 2.99% | Beta (10, 325) |
| Probability of Delirium WEIGHTED | 11.11% | + |
| Probability of Delirium USUAL PRACTICE | 12.37% | Beta (37, 262) |
| Probability of discharge to HOME (delirium) | 49.73% | Dirichlet (2662, 1) |
| Probability of discharge to DEATH (delirium) | 11.17% | Dirichlet (598, 1) |
| Probability of discharge to NURSING HOME (delirium) | 6.59% | Dirichlet (353, 1) |
| Probability of discharge to HOSPITAL (delirium) | 10.42% | Dirichlet (558, 1) |
| Probability of discharge to REHAB (delirium) | 22.08% | Dirichlet (1182, 1) |
| Probability 30-day re-admission (delirium) | 41.18% | Beta (7, 10) |
| Probability of discharge to HOME (no delirium) | 91.66% | Dirichlet (19903, 1) |
| Probability of discharge to DEATH (no delirium) | 0.29% | Dirichlet (64, 1) |
| Probability of discharge to NURSING HOME (no delirium) | 1.00% | Dirichlet (218, 1) |
| Probability of discharge to HOSPITAL (no delirium) | 1.96% | Dirichlet (425, 1) |
| Probability of discharge to REHAB (no delirium) | 5.08% | Dirichlet (1103, 1) |
| Probability 30-day re-admission (no delirium) | 11.01% | Beta (48, 388) |

+ Estimated by ('Probability of Delirium AT Risk' multiplied by 'Probability of AT risk designation') + ('Probability of Delirium LOW Risk' multiplied by (1- 'Probability of AT risk designation'))

